# Impact of CoronaVac on Covid-19 outcomes of elderly adults in a large and socially unequal Brazilian city: A target trial emulation study

**DOI:** 10.1101/2023.06.06.23291015

**Authors:** Higor S. Monteiro, Antonio S. Lima Neto, Rebecca Kahn, Geziel S. Sousa, Humberto A. Carmona, José S. Andrade, Marcia C. Castro

## Abstract

**Background:** Although CoronaVac was the only Covid-19 vaccine adopted in the first months of the Brazilian vaccination campaign, randomized clinical trials to evaluate its efficacy in elderly adults were limited. In this study, we use routinely collected surveillance and SARS-CoV-2 vaccination and testing data comprising the population of the fifth largest city of Brazil to evaluate the effectiveness of CoronaVac in adults 60+ years old against severe outcomes.

**Methods:** Using large observational databases on vaccination and surveillance data from the city of Fortaleza, Brazil, we defined a retrospective cohort including 324,302 eligible adults aged ≥ 60 years to evaluate the effectiveness of the CoronaVac vaccine. The cohort included individuals vaccinated between January 21, 2021, and August 31, 2021, who were matched with unvaccinated persons at the time of rollout following a 1:1 ratio according to baseline covariates of age, sex, and Human Development Index of the neighborhood of residence. Only Covid-19-related severe outcomes were included in the analysis: hospitalization, ICU admission, and death. Vaccine effectiveness for each outcome was calculated by using the risk ratio between the two groups, with the risk obtained by the Kaplan-Meier estimator.

**Results:** We obtained 62,643 matched pairs for assessing the effectiveness of the two-dose regimen of CoronaVac. The demographic profile of the matched population was statistically representative of the population of Fortaleza. Using the cumulative incidence as the risk associated with each group, starting at day 14 since the receipt of the second dose, we found an 82.3% (95% CI 66.3 - 93.9) effectiveness against Covid-19-related death, 68.4% (95% CI 42.3 - 86.4) against ICU admission, and 55.8% (95% CI 42.7 - 68.3) against hospital admission.

**Conclusions:** Our results show that, despite critical delays in vaccine delivery and limited evidence in efficacy trial estimates, CoronaVac contributed to preventing deaths and severe morbidity due to Covid-19 in elderly adults.

## INTRODUCTION

The fast development of vaccines against Covid-19 was unprecedented [1]. However, the emergence and spread of new variants of concern (VOC) [2–6] contributed to new waves of the Covid-19 pandemic worldwide. In Brazil, the Gamma variant, P.1 lineage [7], was regarded as the main etiologic agent that contributed to the pandemic wave in late 2020 and the beginning of 2021 [8], as decreased neutralization activity was observed in previously infected populations [5, 9]. At that time, several potentially safe and efficacious vaccine candidates [10–12] were available. Despite concerns about the possibility of escape from neutralizing antibody responses and loss of vaccine efficacy, results from trials suggested that such vaccines could offer protection to the population and enhance pandemic mitigation strategies [13].

As efficacious vaccines became available, global SARS-CoV-2 mitigation required timely and equitable distribution and uptake worldwide [14, 15]. Nevertheless, despite the rapid development of Covid-19 vaccines, logistic aspects ranging from procurement arrangements to distribution of vaccines resulted in unequal access, disproportionally lower in low- and middle-income countries [16–19]. In Brazil, mRNA-based vaccines were acquired late, and the first vaccines available to the population had inactivated virus technology, such as CoronaVac [20].

Initially, about six million doses of the CoronaVac vaccine were available nationally [21]. By mid to the end of January 2021, Brazilian states’ authorities implemented local mass vaccination campaigns, with the first dose administered on the 18^th^ of January. The campaigns prioritized high-risk groups, starting with health professionals and elderly adults (initially recommended to those aged ≥ 80 years [22]). Despite the legal approval of CoronaVac by national sanitary authorities, significant hesitancy from the federal government resulted in conflicts between the federal and state spheres. The presumed lack of confidence in the efficacy of CoronaVac was somehow reinforced by the limited evidence of clinical trial reports for the Brazilian scenario [23]. Before the start of the national vaccination campaign, one randomized clinical study was carried out between July and December 2020 to estimate CoronaVac’s efficacy in Brazil [24]. The study focused on healthcare professionals and suggested an efficacy estimation of 50.7% (95% CI 36.0-62.0), slightly higher than the minimum required for vaccine approval. As of September 2022, the study has not been peer-reviewed.

Despite limited studies for CoronaVac in Brazil, randomized trials performed in Asian countries reported optimistic results. A phase 3 trial performed in Turkey reported efficacy against confirmed symptomatic disease as 83.5% (95% CI 65.4–92.1) considering a population of adults aged 18-59 years [12]. A similar trial performed in Indonesia revealed an efficacy of 65.3% against symptomatic confirmed infection by SARS-CoV-2, also in the 18-59 years group [25]. Due to the limitations of those studies [26], results from trials carried out in Asian countries might not directly translate to the Brazilian setting. For example, in many Brazilian cities, CoronaVac was the only vaccine available for full immunization among elderly adults in the first three months of the Gamma epidemic wave. In such circumstances, efficacy estimated for young adults offered little valuable information on the protection of adults aged 60 years or more. Vaccine effectiveness studies from observational data offer the opportunity to evaluate the effects of vaccines as they are implemented in noncontrolled settings while accounting for potential sources of bias [27–36]. These analyses have been frequently performed by using traditional study designs, either through test-negative or population-based methods [27, 37–39].

For Brazil, a population-based analysis obtained significant estimates for the effectiveness of CoronaVac against Covid-19 hospitalization, 71.4% (95% CI 70.0-72.4), and Covid-19-related death, 73.7% (95% CI 72.1-75.2) [39]. Using the test-negative case-control design, Ranzani *et al.* [28] have shown that a two-dose regimen of CoronaVac was directly associated with a reduction in symptomatic infection in Brazilian adults aged 70+ years while reporting effectiveness estimates of 55% (95% CI 46.5-62.9) against hospital admission, and 61.2% (95% CI 48.9-70.5) against death, both 14 days after the receipt of the second dose. Other studies were performed using similar standards considering different settings but exhibiting consistent conclusions on the effectiveness of CoronaVac [31, 32].

Here we use a target trial emulation method [30, 40] to estimate the effectiveness of CoronaVac against severe Covid-19 related outcomes in Fortaleza, the fifth largest city of Brazil, during the Gamma wave. The target trial emulation method is rather suitable for observational studies on comparative effectiveness where randomized clinical trials are either missing or provide limited evidence, and when large observational databases are available [40]. To the best of our knowledge, this is the first time that the target trial emulation method is applied to data from vaccination with CoronaVac. We obtained estimates of vaccine effectiveness against hospitalization due to a Covid-19 infection, ICU admission, and Covid-19-related death from the start of the local vaccination campaign in January 2021 until the end of August 2021, when other vaccines were also included.

## MATERIAL AND METHODS

### Study setting

Fortaleza is the fifth largest city in Brazil, the capital of the state of Ceará. Fortaleza had approximately 2.7 million inhabitants (about 360,000 individuals aged 60 years old or more) and a population density of 7,786 inhabitants per km^2^ in 2021 [41]. By August 2022, Fortaleza had already experienced four major Covid-19 epidemic waves, during which 326,065 cases and 11,496 deaths were confirmed, leading to the implementation of two major lockdowns and several physical distancing measures (Fig. 1a).

**Figure 1:**
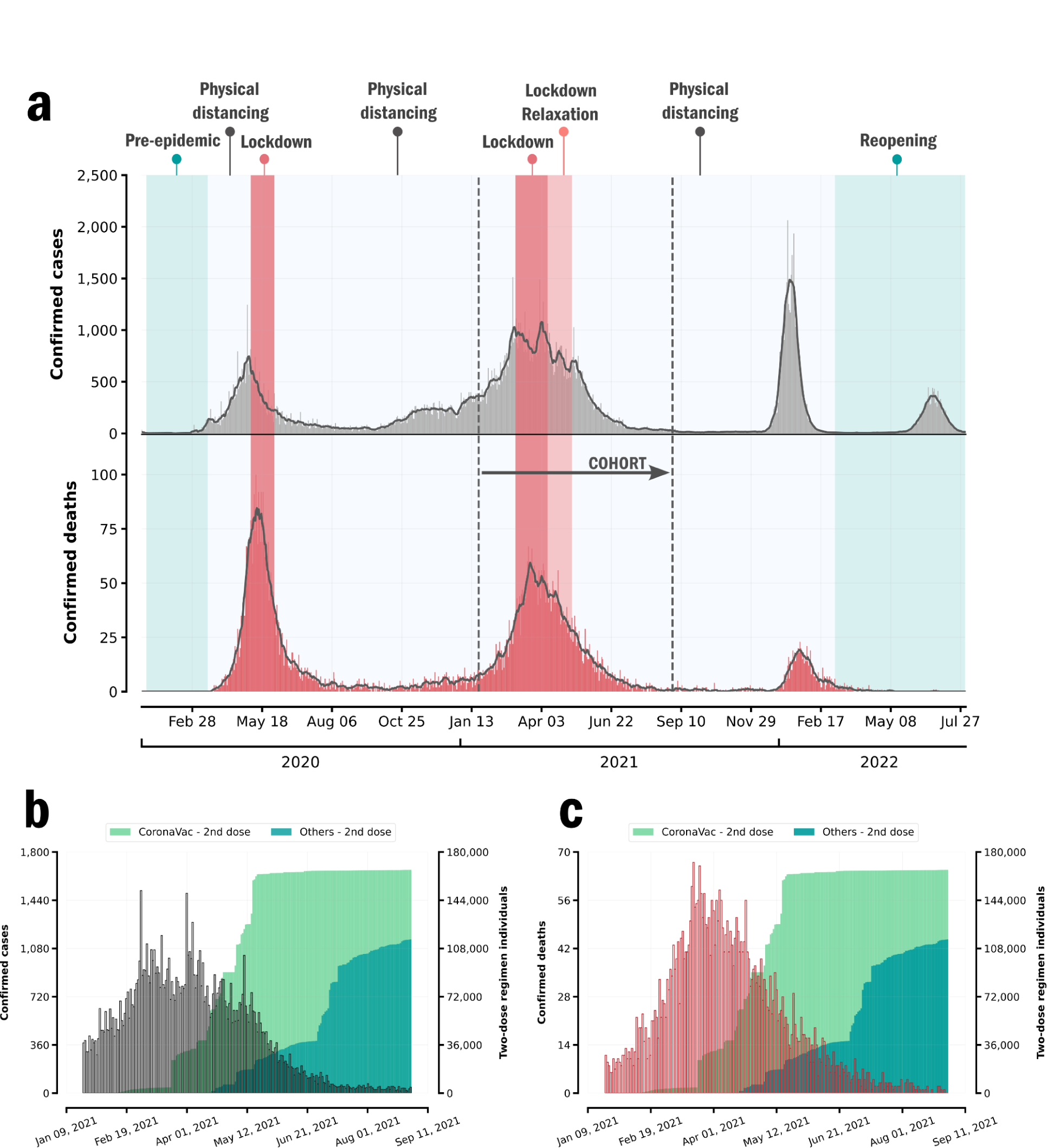
Epidemiology pattern of Covid-19 in Fortaleza, 2020-22. (a) Epidemiological curves of Covid-19 cases and deaths in Fortaleza, and major interventions implemented in the city, Jan 2020-July 2022. (b) Cases (gray) and (c) deaths (red) of Covid-19 and cumulative distribution of individuals vaccinated with 2 doses (we consider a single dose of the Janssen vaccine as equivalent to a two-dose regimen).

Following the national vaccination strategy, the state and municipal health departments of Ceará and Fortaleza, respectively, implemented a mass vaccination campaign, with the first doses administered on January 21, 2021. Following an age prioritization approach, eligibility in the city started with those aged ≥ 75 years and later expanded to those in the age group of 60-75 years [42]. Fortaleza used a two-dose regimen of CoronaVac with an interval of two to four weeks between doses (other vaccines introduced in February used a 12-week interval). At the start of the vaccination campaign, an increasing incidence of confirmed Covid-19 cases was observed, establishing the second major wave in the city, with many cases and deaths occurring during March and April 2021. During the first months of that wave, complete immunization with two doses was possible only for adults receiving CoronaVac, while individuals vaccinated with a different vaccine only started to complete the two-dose regimen on April 6^th^ (Fig. 1b-c).

### Data sources

We obtained individual information on dates of SARS-CoV-2 testing and vaccination from the state registry INTEGRASUS and the municipal registry VACINE JÁ, respectively.

INTEGRASUS is a general surveillance tool created by the State Health Department to integrate epidemiological, hospital, finance, and public management information [43]. Results of all SARS-CoV-2 tests performed in the public and private healthcare networks of the state are stored in the INTEGRASUS registry. VACINE JÁ is a platform to register citizens willing to receive a Covid-19 vaccine and to schedule vaccination dates for registered individuals based on prioritization guidelines. Information on each applied dose, together with personal information on the person receiving the vaccine, is stored in the registry. By January 20 of 2022, almost 2.3 million individuals were registered in VACINE JÁ, representing about 85% of the projected population for Fortaleza in 2021. Also, more than 4.6 million doses were administered, with coverage of *>*90% among the eligible population. Information regarding Covid-19-related outcomes, such as hospitalization, admission to ICU, and death is stored in the national surveillance database for severe acute respiratory illness, SIVEP-Gripe. Through the Brazilian universal health system (SUS) all individuals have access to the public health system, and notification of suspected and confirmed cases and outcomes of Covid-19 is compulsory.

This study was approved by the Ethical Review Board of the Health Department of Fortaleza, process number (CAAE) 63580522.0.0000.0203.

### Study design

In this study, we aimed to emulate a target clinical trial [30, 35, 40] using observational data to evaluate the comparative effectiveness of the two-dose regimen of the CoronaVac vaccine in a population of elderly adults aged ≥ 60 years. Eligible individuals should not have a previous SARS-CoV-2 infection record before the beginning of the study, or should not work as a health professional. For each individual, the eligibility criterion on previous SARS-CoV-2 infection was verified by checking the existence of any positive SARS-CoV-2 RT-PCR or rapid antigen test, or notification of first symptoms or Covid-19-related outcome dated before the start of the study.

Health professionals, including frontline health workers, clinicians, and administrative staff, were excluded from the cohort because of their high variability in exposure. Finally, we also excluded any individual missing essential demographic information, without documented residency in Fortaleza, and anyone with irregular dates either on vaccination or severe outcomes.

We defined a rolling retrospective cohort starting on the first day of the vaccination campaign, January 21, 2021, and ending on August 31, 2021. Individuals vaccinated with CoronaVac and their respective controls were selected during this period, while satisfying the eligibility criteria on the day of dose receipt of the vaccinated person. Individuals were matched with controls dynamically as the cohort proceeded to mimic the vaccine rollout as occurred in the real-world setting. In the analysis for the two-dose regimen, each individual is recruited on the day of the receipt of the second dose. To control for potential confounding, each person receiving the vaccine is immediately matched with a control based on baseline covariates associated with the chance of being vaccinated and socioeconomic status given by residence location. Variables chosen for matching were age, sex, and the Human Development Index (HDI) of the neighborhood of residence [44]. Matching by age considered the age of the unvaccinated control to be within the interval [*x−δ, x*+*δ*], where *x* represents the age of the vaccinated person and *δ* = 1 year. Matching by the HDI was done considering five categories commonly used in the city to distinguish the socioeconomic status of neighborhoods: very low (0.000 to 0.499), low (0.500 to 0.599), medium (0.600 to 0.699), high (0.700 to 0.799) and very high (0.800 to 1.000). Matching was performed using a 1:1 ratio, and matched pairs were followed throughout the cohort for each outcome of interest (Fig. 2).

**Figure 2:**
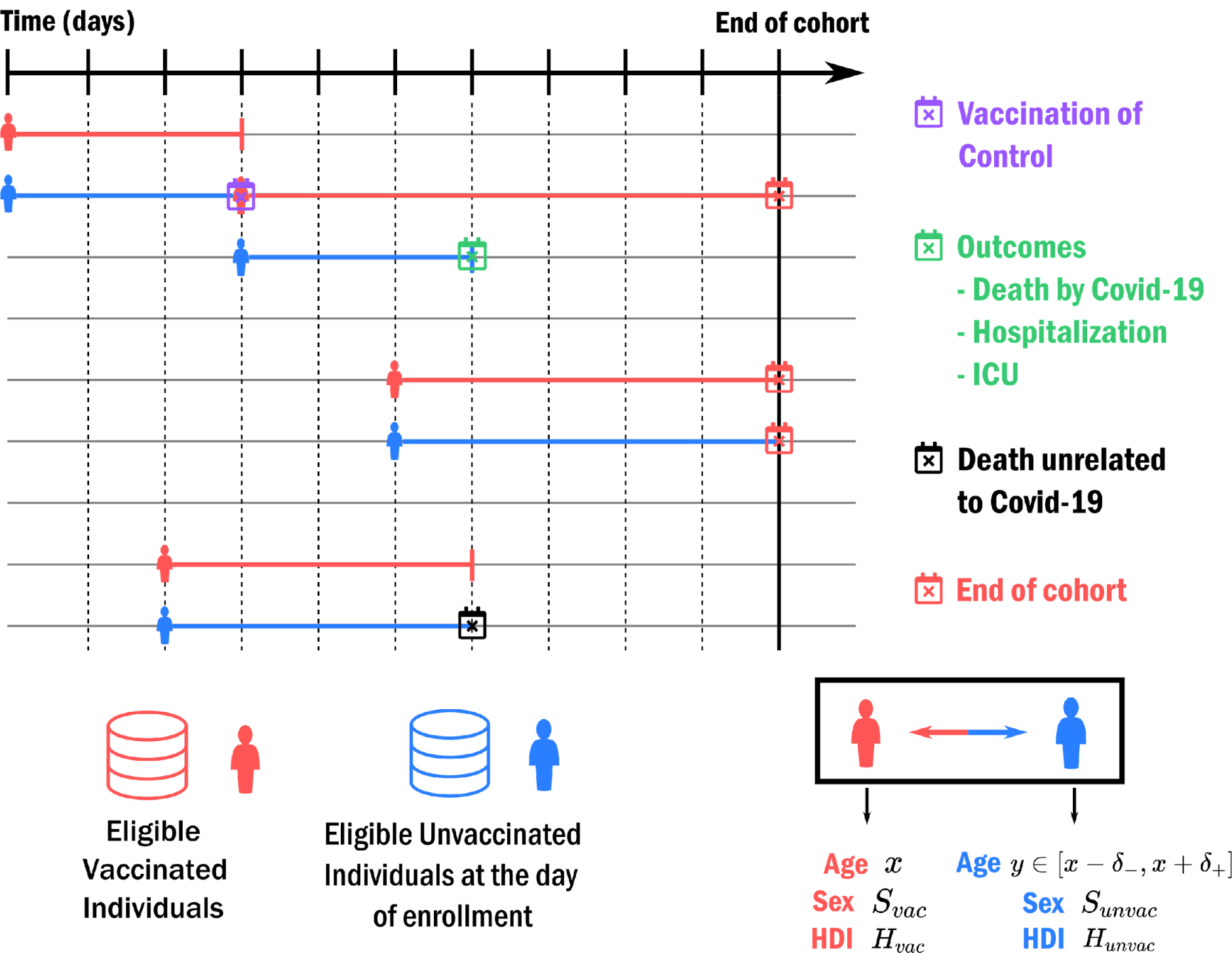
Matching procedure. Each eligible individual vaccinated is matched on the day of vaccine receipt with a current unvaccinated person. Unvaccinated controls are matched to the vaccinated person if they have the same sex, age within at most one year of difference (*δ_−_* = *δ*_+_ = 1 year), and the same HDI of their neighborhood of residence. The baseline covariates for each individual are considered on the day of vaccine receipt of the vaccinated person at the time of matching. For each outcome, matched pairs are followed until either the outcome or a censoring event is observed. The follow-up of a matched pair is censored if either the unvaccinated control receives a vaccine (for which the person is matched with a new unvaccinated in case the vaccine is CoronaVac), death unrelated to Covid-19 of any of the paired individuals or the end of the follow-up, defined here as 90 days after the first day of follow-up.

The outcomes here considered were hospital admission related to Covid-19 confirmed infection, critical Covid-19 represented by cases admitted to ICU, and Covid-19-related deaths. These outcomes have the most relevance for public health since they are directly related to potential healthcare disruption and mortality. Also, such endpoints are less sensitive to bias related to underreporting. For each outcome, matched pairs were followed from the day of receipt of the second dose of the vaccinated individual until either the outcome of interest or a censoring event is observed. Censuring events include the end of the cohort study, a death not related to Covid-19, and vaccination of the matched control. To avoid bias due to outcomes observed during the first days after the application of the second dose, we only considered outcomes that occurred from the 14^th^ day after that dose until the end of the follow-up. Therefore, the first 13 days of the matched pair are considered to be the period where there is no detectable effect from the vaccine.

### Statistical Analysis

We estimated the effectiveness of CoronaVac against Covid-19-related outcomes after 13 days from the application of the second dose. We also estimated effectiveness during the periods 0-13 and ≥ 14 days for a single dose for further sensitivity analysis. After matching individuals, we performed survival analysis for both vaccinated and unvaccinated groups considering the defined cohort period. Survival curves and cumulative incidence curves for both groups were obtained with the nonparametric Kaplan-Meier estimator [45]. For each outcome, we estimated the risk ratio using only the matched pairs in which both individuals were still at risk 13 days after the receipt of the vaccine (date for the vaccinated group). For the hospitalization outcome, we also provided stratified estimates considering specific age groups and sex.

Considering the observational nature of the study, bias due to unmeasured confounding is possible. Although the matching procedure represents a robust adjustment, complete control for confounding is not certain in observational studies. Here, confounding may exist if vaccinated and unvaccinated groups differ in characteristics not observed. To improve the statistical quality of the matched pairs, we repeated the matching process with different random seeds to obtain several different configurations of controls for the same vaccinated group. For each realization of the matching process, we obtained estimates of vaccine effectiveness, and the 95% CIs were calculated using the nonparametric percentile bootstrap method with 1,000 repetitions. Therefore, we evaluated different estimations of the effectiveness and their fluctuations within different realizations. Vaccine effectiveness was estimated as one minus the risk ratio. We have excluded estimates for analyses where less than 10 observations of an outcome were observed across the two groups.

To verify potential unmeasured differences between vaccinated and unvaccinated groups, we considered the period immediately after the application of the first dose until day 13 to compare the cumulative incidence curves between groups. This period was excluded from the estimation of effectiveness because it is expected that the risks of both groups are equivalent (no effect from the vaccine was observed in that period). Therefore, the risk ratio between groups should be close to one during this interval. We also obtained the risk ratio for this period as an average from different configurations of controls to reduce the influence of fluctuations for scenarios where there is a small number of outcomes. In addition, a sensitivity analysis was conducted to evaluate the potential for selection bias due to informative censoring. In this analysis, controls who were subsequently vaccinated were censored only after 6 and 13 days, where the effects of vaccination are expected to be relevant. Analyses were performed in Python, version ≥3.9.

## RESULTS

### Study population

Among the 2,275,309 individuals included in the VACINE JÁ database, 1,693,174 were vaccinated during the duration of the cohort study. Four types of vaccines were administered during the period of the cohort, and CoronaVac was the most used among elderly adults in the first three months of the campaign. For individuals aged ≥ 60 years, 186,939 were vaccinated with CoronaVac before the end of the study and 324,302 were eligible to participate in the cohort. From the eligible population, 62,482 were matched to unvaccinated controls for the two-dose regimen cohort. For the analysis regarding a single dose of CoronaVac, 124,609 were matched to unvaccinated controls. The total follow-up time was 1,908,853 person-days, with a median follow-up time of 36 days (IQR 24-54) after the first 13 days in the two-dose regimen in both groups, with a maximum follow-up time of 90 days. During the cohort, a total of 149,694 confirmed tests for Covid-19 were performed, consisting of both RT-PCR and rapid antigen tests.

Most of the controls received a first dose of a vaccine during the study (Table 1); 30% of the controls received CoronaVac, and then were included in the vaccinated group and matched with a new unvaccinated control, while 35.9% of the controls received a different vaccine: 30.9% ChAdOx1 nCoV-19 (AstraZeneca), 4.5% BNT162b2 (Pfizer-BioNtech), and 0.6% Ad26.COV2.S (Janssen). A total of 41,157 matched pairs in which controls were eventually vaccinated during the follow-up were censored, and about 10% of the eligible persons were dropped from the analysis due to irregular information regarding either vaccine or outcome dates or due to missing essential data for matching (Fig. 3).

**Figure 3:**
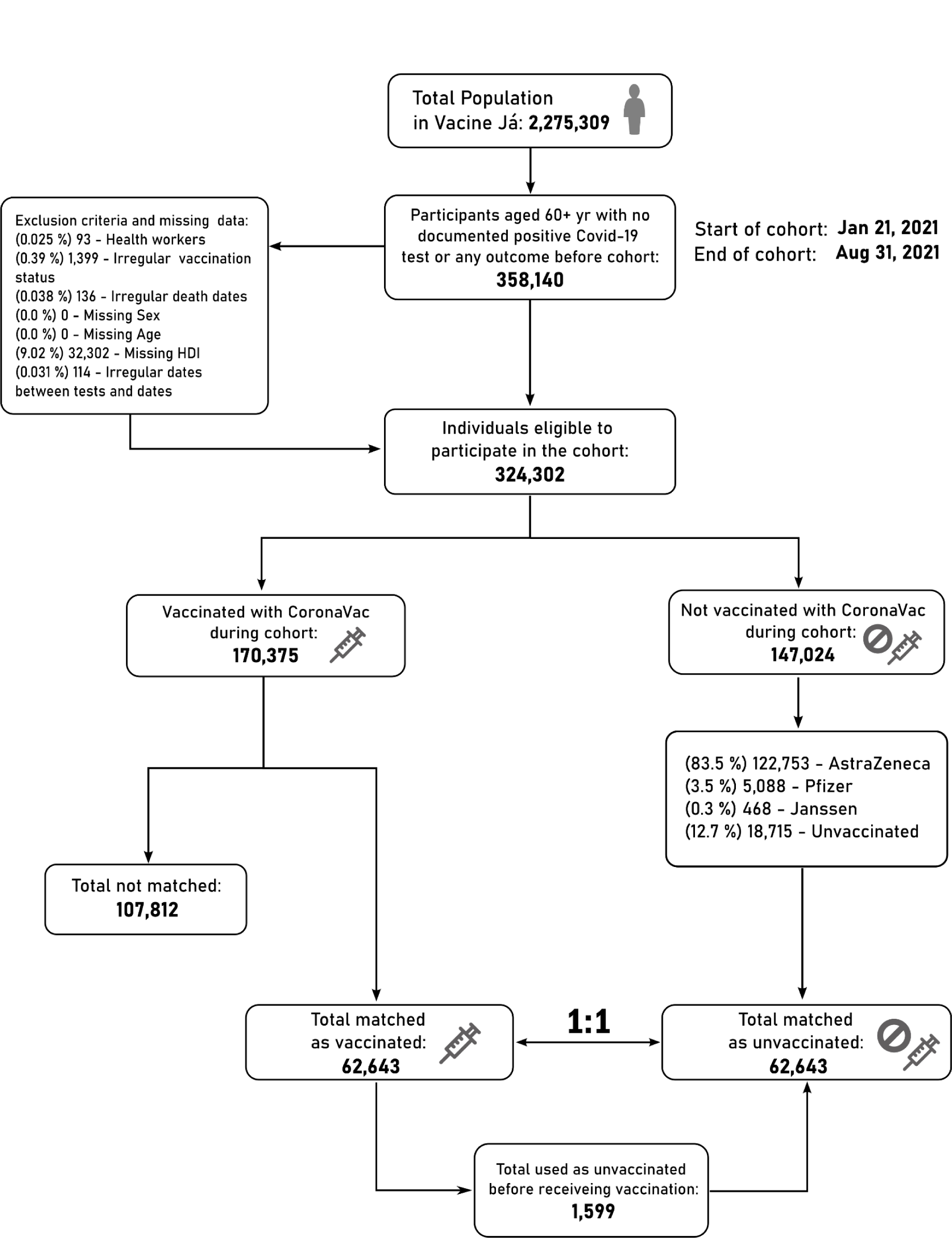
Flow chart of cohort recruitment.

**Table 1:**
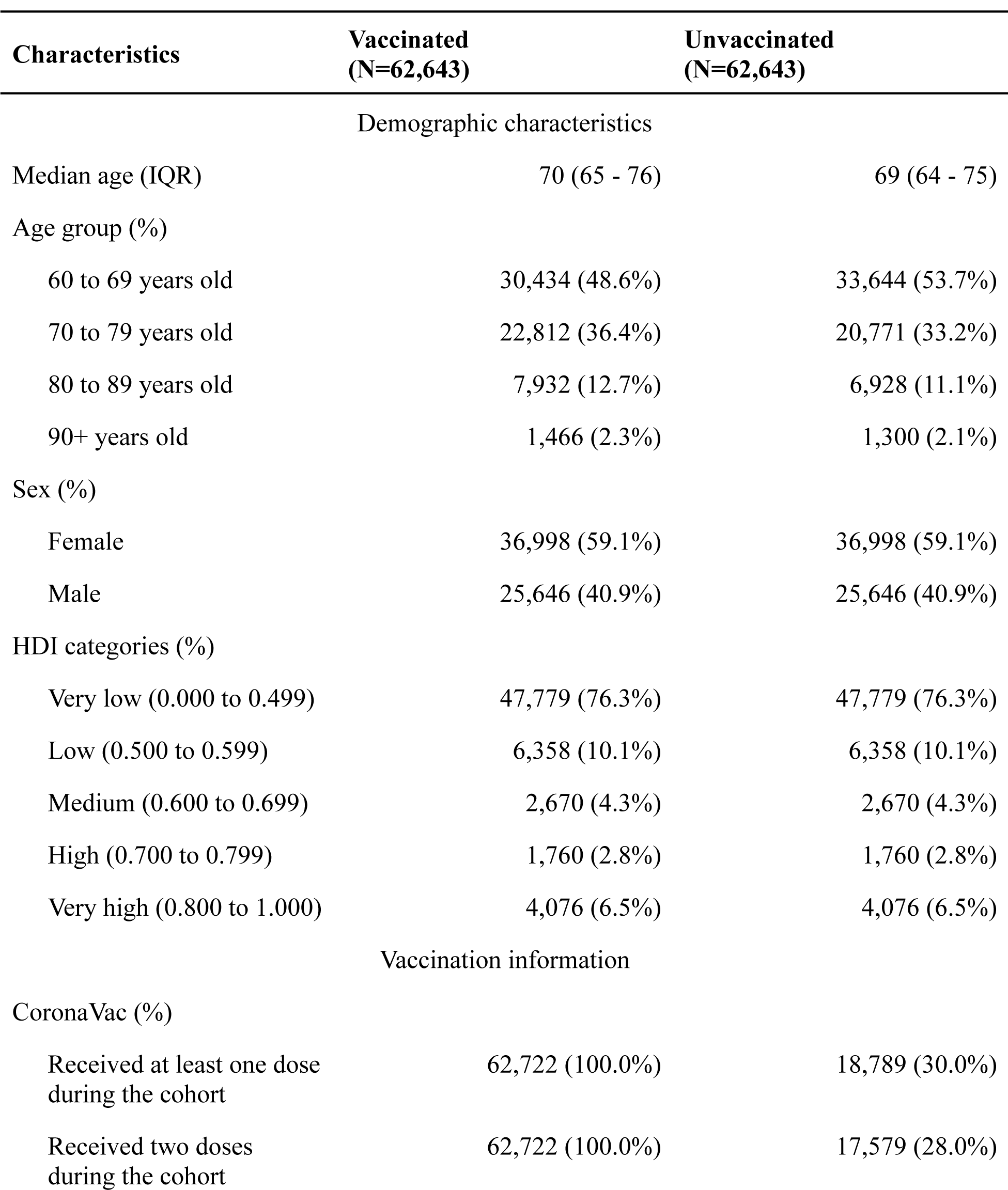
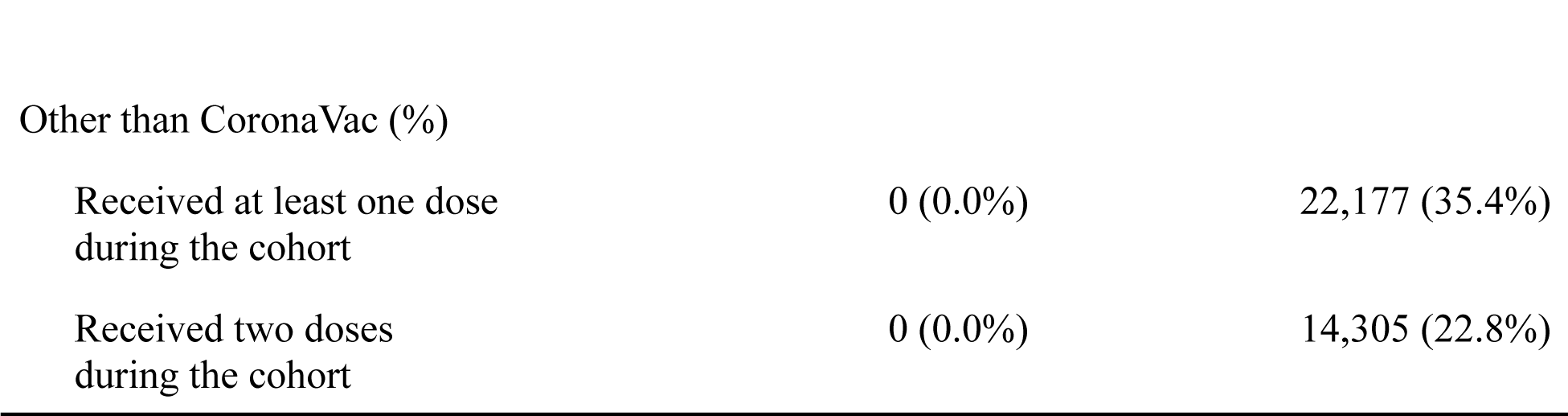
Demographic characteristics and vaccination information of the matched pairs at baseline.

| Characteristics | Vaccinated<br>(N=62,643) | Unvaccinated<br>(N=62,643) |
| --- | --- | --- |
| Demographic characteristics |  |  |
| Median age (IQR) | 70 (65 - 76) | 69 (64 - 75) |
| Age group (%) |  |  |
| 60 to 69 years old | 30,434 (48.6%) | 33,644 (53.7%) |
| 70 to 79 years old | 22,812 (36.4%) | 20,771 (33.2%) |
| 80 to 89 years old | 7,932 (12.7%) | 6,928 (11.1%) |
| 90+ years old | 1,466 (2.3%) | 1,300 (2.1%) |
| Sex (%) |  |  |
| Female | 36,998 (59.1%) | 36,998 (59.1%) |
| Male | 25,646 (40.9%) | 25,646 (40.9%) |
| HDI categories (%) |  |  |
| Very low (0.000 to 0.499) | 47,779 (76.3%) | 47,779 (76.3%) |
| Low (0.500 to 0.599) | 6,358 (10.1%) | 6,358 (10.1%) |
| Medium (0.600 to 0.699) | 2,670 (4.3%) | 2,670 (4.3%) |
| High (0.700 to 0.799) | 1,760 (2.8%) | 1,760 (2.8%) |
| Very high (0.800 to 1.000) | 4,076 (6.5%) | 4,076 (6.5%) |
| Vaccination information |  |  |
| CoronaVac (%) |  |  |
| Received at least one dose during the cohort | 62,722 (100.0%) | 18,789 (30.0%) |
| Received two doses during the cohort | 62,722 (100.0%) | 17,579 (28.0%) |
| Other than CoronaVac (%) |  |  |
| Received at least one dose<br>during the cohort | 0 (0.0%) | 22,177 (35.4%) |
| Received two doses<br>during the cohort | 0 (0.0%) | 14,305 (22.8%) |

### Vaccine Effectiveness

During the duration of the cohort, a total of 4,461 hospitalizations, 1,025 ICU admissions, and 916 deaths were recorded in the eligible population. Considering the population of matched pairs and a mean follow-up of 40.5 days (IQR 24-55), we observed 1,951 documented infections, of which 402 required hospitalizations, 92 were admitted to the ICU and 112 resulted in death. The cumulative incidence curves generated by the Kaplan-Meier estimator are shown in Fig. 4 for each outcome considered. To estimate the vaccine effectiveness for individuals still at risk after 13 days of the second dose, we calculated the risk ratio at day 90 of follow-up. The effectiveness of the two-dose regimen against hospitalization due to Covid-19, compared with the unvaccinated group, was estimated at 55.8% (95% CI 42.7-68.3) with 135 events for the unvaccinated versus 60 events for the vaccinated group with two doses. For critical disease manifestation, represented by the admission to ICU due to Covid-19, the effectiveness was estimated at 68.4% (95% CI 42.3-86.4) with 32 events for the unvaccinated group against 10 events for the vaccinated group with two doses. For Covid-19-related deaths, we estimated that the two-dose regimen of CoronaVac had an 82.3% (95% CI 66.3-93.9) effectiveness, with 46 events for the unvaccinated group against 8 events for the vaccinated group. These estimates, together with the risk in each group, are presented in Table 2. No significant effect was found for a single dose (Fig. S3).

**Figure 4:**
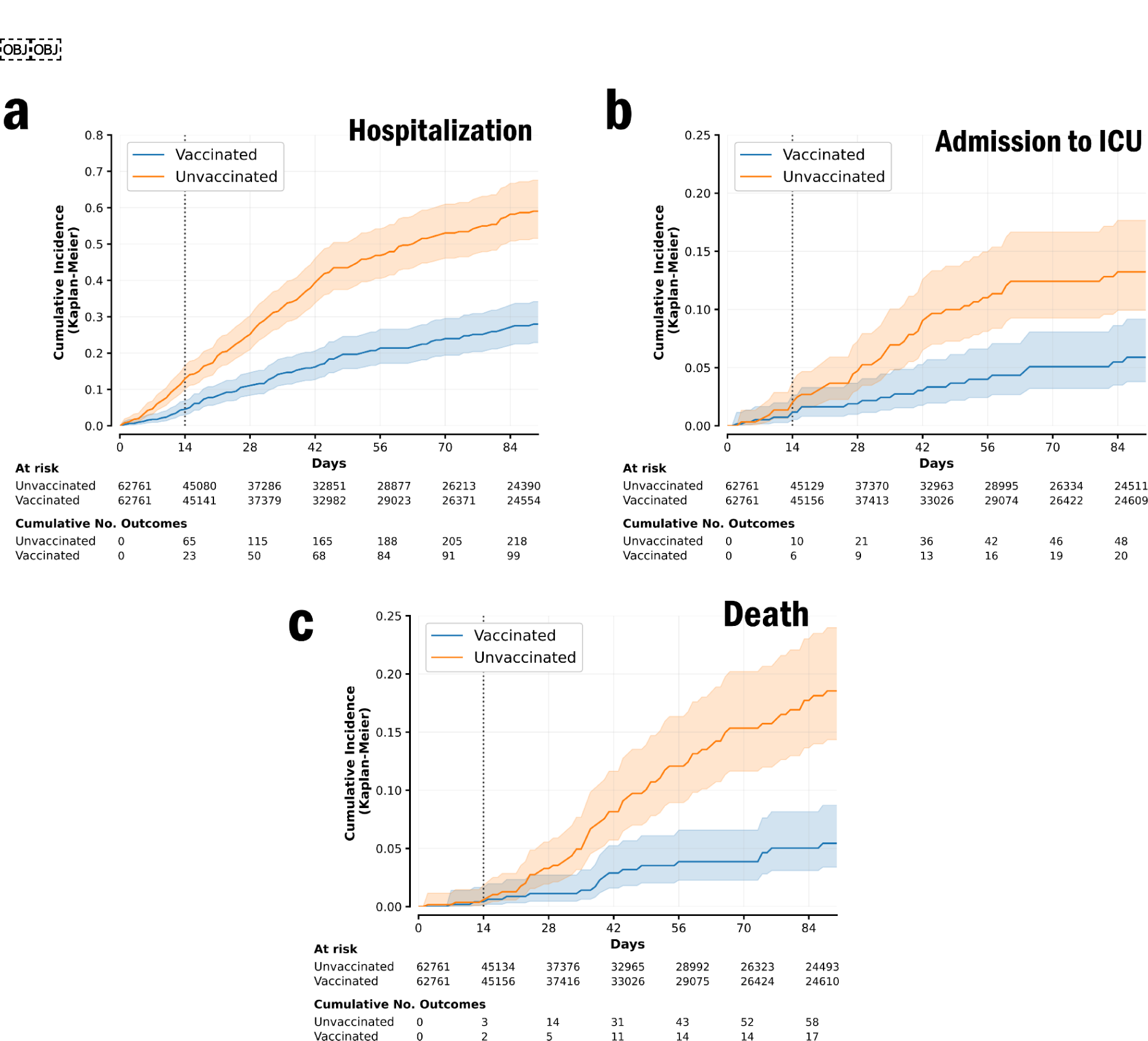
Cumulative incidence curves comparing the risk between individuals vaccinated with two doses of CoronaVac and unvaccinated individuals. Curves were estimated with the Kaplan-Meier estimator for (a) hospital admission, (b) ICU admission, and (c) Covid-19-related death. An early divergence is observed for the outcome of hospitalization. This divergence might be due to the presence of unreported infected individuals before the day of matching. Therefore, to estimate vaccine effectiveness, outcomes were considered only from day 14 of receipt of the second dose for the vaccinated person (dotted line in the incidence curves).

**Table 2:** Effectiveness of the two-dose regimen of CoronaVac against severe outcomes. Number of outcomes and risks are shown for both two-dose regimen individuals and unvaccinated individuals. 95% confidence intervals of the estimates were obtained by using the percentile bootstrap method with 1000 repetitions.

| **Outcome** | **Unvaccinated**<br>**Individuals** |  | **Two-dose regimen**<br>**Individuals** |  | **1 - RR**<br>**(95% CI)** |
| --- | --- | --- | --- | --- | --- |
|  | **Events** | **Risk per**<br>**100,000**<br>**individuals** | **Events** | **Risk per**<br>**100,000**<br>**individuals** |  |
| Hospitalization | 135 | 570.0 | 60 | 251.9 | 55.8% (42.7 - 68.3) |
| Admission to ICU | 32 | 134.6 | 10 | 41.9 | 68.4% (42.3 - 86.4) |
| Death | 46 | 193.7 | 8 | 33.5 | 82.3% (66.3 - 93.9) |

The time for the vaccine to produce a detectable effect is identified by the number of follow-up days at which the risk curves between the vaccinated and unvaccinated groups start to diverge. Among the available outcomes in our study, we used the outcome of hospitalization to estimate such divergence. Fig. S4 shows the distribution of the risk ratios obtained for different configurations of unvaccinated controls on day 14 after the first dose. As observed, the distribution is concentrated close to one, reinforcing the assumption of exchangeability of the matched pairs and the robustness of the matching procedure.

We also performed an effectiveness estimation for hospitalization in subpopulation groups. For more severe outcomes, such as admission to ICU and death, the number of outcomes captured during the follow-up was not large enough to provide robust estimates for any specific strata. For hospitalization, we observed larger vaccine effectiveness in women, 68.5% (95% CI 54.1-80.6), compared to men, 35.2% (95% CI 3.4-60.3). The same analysis was performed by age groups: effectiveness of 69.4% (95% CI 47.1-87.1) for the 60-69 group, 47.9% (95% CI 19.5-70.0) for individuals in the 70-79 group, and finally 54.2% (95% CI 30.0-74.0) for individuals aged 80+ years. In Table 3 we present the vaccine effectiveness and risk values obtained from the subgroups’ analysis on hospitalization.

**Table 3:** Subpopulation analysis of the effectiveness of two-dose regimen of CoronaVac for the outcome of hospital admission. 95% confidence intervals of the estimates were obtained by using the percentile bootstrap method with 1000 repetitions.

| Subgroup | Unvaccinated<br>Individuals |  | Two-dose regimen<br>Individuals |  | 1 - RR (95% CI) |
| --- | --- | --- | --- | --- | --- |
|  | Hospital<br>Admissions | Risk per<br>100,000<br>individuals | Hospital<br>Admissions | Risk per<br>100,000<br>individuals |  |
| Sex |  |  |  |  |  |
| Male | 52 | 491.4 | 33 | 310.6 | 35.2% (3.4 - 60.3) |
| Female | 83 | 630.7 | 27 | 204.4 | 68.5% (54.1 - 80.6) |
| Age (years) |  |  |  |  |  |
| 60 to 69 | 38 | 309.8 | 11 | 89.4 | 69.4% (47.1-87.1) |
| 70 to 79 | 49 | 592.9 | 26 | 312.8 | 47.9% (19.5-70.0) |
| 80+ | 47 | 1452.4 | 23 | 704.4 | 54.2% (30.0 - 74.0) |

To evaluate the consistency of our estimates, we conducted a series of sensitivity analyses regarding the matching procedure and follow-up rules. First, we create different samples of unvaccinated controls to evaluate the robustness of the matching procedure. The different estimates of vaccine effectiveness obtained for each random seed (Fig. S5) display a consistent behavior compared with the estimations from the original cohort. Given each different configuration, we can directly assess the variability of effectiveness within the target population between different samples. Second, we delay the timing of censorship for matched pairs where the control individual receives the vaccine. In such cases, no effect from the vaccine is considered in the first days after receipt of the first dose, and thus we allowed outcomes to be recorded if they occurred during this period for the matched pair. We considered both a delay of 6 and 13 days after the receipt of the first dose for the control individual. Results indicate the consistency between estimates of vaccine effectiveness since a very small variation is observed within the same control configuration (Table S1).

## DISCUSSION

We estimated the effectiveness of the two-dose regimen of the inactivated SARS-CoV-2 vaccine CoronaVac and found it to be effective against severe Covid-19-related outcomes in elderly adults. The study was performed in the fifth largest city of Brazil considering individuals aged 60+ years old during an epidemic wave associated with the Gamma variant. Different from other studies implemented in Brazil, our study followed the framework of a target trial emulation by using a large retrospective cohort from a populous city. The effectiveness of the two-dose regimen of CoronaVac, compared with unvaccinated individuals, was estimated to be 55.8% against a hospital admission, 68.4% against an ICU admission, and 82.3% in preventing a Covid-19-related death.

The two-dose regimen of CoronaVac displayed large protection against hospitalization in the female population (68.5%). Although effectiveness for the male population was lower (35.2%) we note the wide confidence interval and smaller sample size for this subgroup, which does not allow us to contrast effectiveness between both groups. Considering different groups of age, CoronaVac exhibited higher protection against hospital admission among individuals aged 60 to 69 years old (69.4%) while it displayed reduced, albeit significant, effectiveness for older individuals: 47.9% for the 70-79 years group and 54.2% for individuals aged 80+ years. Wide confidence intervals in the estimates for the 70-79 years group and the 80+ years group reveal uncertainties that do not allow a proper comparison between the effectiveness of the two groups. Nevertheless, we observe in Table 3 that both unvaccinated and vaccinated individuals aged 80+ years have a significantly higher risk of hospital admission compared to younger individuals in the study. This pattern of high-risk groups is consistent with the prioritization by age for the vaccination schedule implemented in Fortaleza and across other Brazilian cities. The sample size of our study did not allow for the same subgroup analysis considering the outcomes of ICU admission and death.

Due to the shorter interval between CoronaVac doses (two to four weeks), compared to other vaccines implemented during the recruitment period (7 months), we focused our analysis on individuals with a complete vaccination regimen (two doses of CoronaVac). Although a considerable percentage of eligible individuals followed the official vaccination schedule, a comparable percentage did not (Fig. S1). This irregularity is not only due to imperfect adherence by the population, but also with issues of vaccine supply during the cohort period. Therefore, we also estimated the risk for individuals under a single dose regimen of CoronaVac considering the same severe outcomes mentioned before. There were no significant differences between individuals with a single dose and unvaccinated individuals during the first weeks of vaccine receipt (Fig. 3), a result consistent with analysis provided in other studies and reports from clinical trials [12, 28].

Compared with other observational studies on the effectiveness of CoronaVac in Brazil, our analysis provides comparable results and new insights into the national vaccination campaign. A study that investigated adults aged 70+ years during a gamma variant [28] found similar results regarding the effectiveness of CoronaVac against hospital admission due to Covid-19, but lower effectiveness against a Covid-19-related death. The disagreement, however, was most likely due to the presence of a younger population (60 to 69 years) in our study given the age mortality pattern of Covid-19 [46]. Therefore, divergences in effectiveness estimates may be associated with two crucial characteristics of our study. First, we considered the particular setting of a highly populous city and included individuals aged 60 years or older. Second, we have adopted the target trial framework, which largely differs from the methodologies employed in other observational studies done for CoronaVac. In this sense, our study more closely approximates the set of studies that evaluated the effectiveness of the BNT162b2 mRNA vaccine in high-income countries [30,35].

The direct emulation of a target trial through a retrospective cohort provides a range of advantages over other study designs [40], but demands a set of stricter requirements for its successful implementation. To include essential eligibility criteria and to enforce exchangeability between vaccinated individuals and unvaccinated controls at the baseline of the cohort, the availability of detailed information on individuals, ranging from demographic data to clinical records, is indispensable. Although Brazil has a universal health system, with information systems gathering standardized information collected nationwide, detailed individual data also exist in different information systems across states. Fortaleza has local registries comprising nearly the entire population of elderly adults in Fortaleza, which allowed us to select a homogeneous eligible population and to set up a robust matching procedure despite the lack of clinical variables for each individual. To enforce exchangeability between compared groups, we matched vaccinated and unvaccinated individuals based on age, sex, and HDI of the neighborhood of the residence. The use of the HDI was essential to uniformize the matched population without drastically reducing the final sample because Fortaleza is a highly unequal city [47].

As the registration on the platform VACINE JÁ is done by individuals willing to receive a vaccine, there is a valid concern that the population associated with the registry might not represent well the population of Fortaleza. However, while it is expected that specific groups should be excluded from the database, such as individuals who are avowedly against vaccines, the uptake in Fortaleza was very high, reinforced by the obligation of “vaccination passports” [48]. Indeed, the main characteristics of the 60+ years population registered in VACINE JÁ and the estimated population for Fortaleza in 2021 are quite similar (Fig. S2). Therefore, we were able to build a representative cohort including a large portion of the eligible population.

Nevertheless, residual and unmeasured confounding remain a limitation in our study considering its observational nature, and it arises mainly from unmeasured differences between the vaccinated individuals and unvaccinated controls. Although the procedure of matching vaccinated and unvaccinated individuals was adopted to mitigate such confounding, the lack of clinical variables, such as the existence of comorbidities, could affect our results with residual confounding. We verified the consistency of the matched pairs by first creating equivalent cohorts with different samples of unvaccinated controls, drawn from the eligible population, and then matching them with the set of vaccinated individuals. For each sample, we estimated the risk ratio of hospital admission between the vaccinated and controls after 14 days of the receipt of the first dose to obtain their distribution (Fig. S4). The risk ratio distribution of the different cohorts is centered around one, suggesting robust exchangeability of both groups at baseline. Here, we used the outcome of hospital admission as a close proxy to the symptomatic disease outcome, but with less sensitivity to bias due to health-seeking behavior. To avoid additional residual confounding expected to arise from specific groups with high variability in their exposure, such as health professionals (e.g., health workers, clinicians, and administrative staff), the eligibility criteria used to build the cohort did not include them.

The use of several cohorts with different configurations of unvaccinated controls enabled a more structured evaluation of the uncertainties in the analysis. While estimates of vaccine effectiveness were similar for the outcomes of hospital admission and a Covid-19-related death, much larger variability was observed in the estimates for the outcome of admission to the ICU (Fig. S5). This sensitivity analysis shed light on the effectiveness obtained for the subpopulation results provided in Table 3, where we observed wider confidence intervals (Fig. S6-S7). Despite those uncertainties, the distribution of effectiveness for both stratifications is concentrated around their respective average values. Therefore, this sensitivity analysis not only corroborates the patterns discerned in Table 2-3 but also provides further insights into their uncertainties.

Finally, we addressed the potential selection bias that might arise from informative censoring. We considered the scenario where controls might be infected but do not present significant symptoms on the day of vaccine receipt (if the control receives a dose of any Covid-19 vaccine), and therefore we delayed by 6 and 13 days the censoring of the respective matched pair. Results show similar estimates (Table S1) compared to those shown in Table 2, suggesting small interference of selection bias in the estimates.

At the time of writing, the efficacy of CoronaVac for elderly adults remained unresolved with limited clinical evidence. In Brazil, a clinical trial performed on the population of young health professionals [24] in 2020 has not yet been peer-reviewed and published. Fueled by misinformation from the federal government, a negative reputation for CoronaVac during the Brazilian mass vaccination was developed, causing the interruption of its production due to a lack of demand in 2022. Nevertheless, CoronaVac was the only available vaccine providing complete immunization through a two-dose regimen for elderly adults for 3 months, in the worst moment of the pandemic in Brazil due to the Gamma variant. Our study, grounded in a novel methodology, estimated the high effectiveness of CoronaVac against severe outcomes, especially against the critical outcomes of ICU admission and death related to Covid-19. Our analysis suggests that despite the critical delay in the delivery of immunization, CoronaVac played a crucial role in preventing an even more dramatic scenario in terms of lives lost to Covid-19 [49].

## Data Availability

The datasets generated and/or analyzed during the current study are available in the GitHub repository together with associated code: https://github.com/mcastrolab/coronavac-ve-brazil.

## ACKNOWLEDGEMENTS

This study was funded by X, grant number X. The funder played no role in study design, data collection, analysis and interpretation of data, or the writing of this manuscript.

## REFERENCES

[1] Ball P. The lightning-fast quest for COVID vaccines — and what it means for other diseases. Nature. 2020.

[2] Krause PR, Fleming TR, Longini IM, Peto R, Briand S, Heymann DL, et al. SARS-CoV-2 Variants and Vaccines. New England Journal of Medicine. 2021;385(2):179–86

[3] Krause PR, Fleming TR, Peto R, Longini IM, Figueroa JP, Sterne JAC, et al. Considerations in boosting COVID-19 vaccine immune responses. The Lancet. 2021;398(10308):1377–80.

[4] Tao K, Tzou PL, Nouhin J, Gupta RK, de Oliveira T, Kosakovsky Pond SL, et al. The biological and clinical significance of emerging SARS-CoV-2 variants. Nature Reviews Genetics. 2021;22(12):757–73.

[5] Sabino EC, Buss LF, Carvalho MPS, Prete Jr CA, Crispim MAE, Fraiji NA, et al. Resurgence of COVID-19 in Manaus, Brazil, despite high seroprevalence. The Lancet. 2021;397(10273):452–5.

[6] Lemey P, Ruktanonchai N, Hong S, Colizza V, Poletto C, Van den Broeck F, et al. SARS-CoV-2 European resurgence foretold: interplay of introductions and persistence by leveraging genomic and mobility data. Res Sq [Preprint]. 2021.

[7] Faria NR, Mellan TA, Whittaker C, Claro IM, da S Candido D, Mishra S, et al. Genomics and epidemiology of the P.1 SARS-CoV-2 lineage in Manaus, Brazil. Science. 2021;372(6544):815-21.

[8] Buss LF, Prete CA, Abrahim CMM, Mendrone A, Salomon T, de Almeida-Neto C, et al. Three-quarters attack rate of SARS-CoV-2 in the Brazilian Amazon during a largely unmitigated epidemic. Science. 2021;371(6526):288-92.

[9] The Lancet. COVID-19 in Latin America: emergency and opportunity. The Lancet. 2021;398(10295):93.

[10] Polack FP, Thomas SJ, Kitchin N, Absalon J, Gurtman A, Lockhart S, et al. Safety and Efficacy of the BNT162b2 mRNA Covid-19 Vaccine. New England Journal of Medicine. 2020;383(27):2603–15.

[11] Voysey M, Clemens SAC, Madhi SA, Weckx LY, Folegatti PM, Aley PK, et al. Safety and efficacy of the ChAdOx1 nCoV-19 vaccine (AZD1222) against SARS-CoV-2: an interim analysis of four randomised controlled trials in Brazil, South Africa, and the UK. The Lancet. 2021;397(10269):99–111.

[12] Tanriover MD, Doğanay HL, Akova M, Güner HR, Azap A, Akhan S, et al. Efficacy and safety of an inactivated whole-virion SARS-CoV-2 vaccine (CoronaVac): interim results of a double-blind, randomised, placebo-controlled, phase 3 trial in Turkey. The Lancet. 2021;398(10296):213–22.

[13] Thye AYK, Law JWF, Pusparajah P, Letchumanan V, Chan KG, Lee LH. Emerging SARS-CoV-2 Variants of Concern (VOCs): An Impending Global Crisis. Biomedicines. 2021;9(10).

[14] Emanuel EJ, Persad G, Kern A, Buchanan A, Fabre C, Halliday D, et al. An ethical framework for global vaccine allocation. Science. 2020;369(6509):1309-12.

15. World Health Organization. A Global Framework to Ensure Equitable and Fair Allocation of COVID-19 Products and Potential implications for COVID-19 Vaccines; 2020.

[16] Hassan F, London L, Gonsalves G. Unequal global vaccine coverage is at the heart of the current covid-19 crisis. BMJ. 2021;375. n3074

[17] Boeck KD, Decouttere C, Vandaele N. Vaccine distribution chains in low- and middle-income countries: A literature review. Omega. 2020;97:102097.

[18] da Fonseca EM, Shadlen KC, Bastos FI. The politics of COVID-19 vaccination in middle-income countries: Lessons from Brazil. Social Science Medicine. 2021;281:114093.

[19] Usher AD. A beautiful idea: how COVAX has fallen short. The Lancet. 2021;397(10292):2322–5.

20. Innovations in Healthcare. The Curious case of upper-middle income countries. 2021. Available from: https://www.innovationsinhealthcare.org/the-curious-case-of-upper-middle-income-countries/.

21. Agência Senado. Brasil poderia ter sido primeiro do mundo a vacinar, afirma Dimas Covas à CPI. 2021. Available from: https://www12.senado.leg.br/noticias/materias/2021/05/27/brasil-poderia-ter-sido-primeiro-do-mundo-a-vacinar-afirma-dimas-covas-a-cpi.

22. Ministério da Saúde. Plano Nacional de Operacionalização da Vacinação Contra a Covid-19. Brasília, BR; 2021.

23. Folha de São Paulo. Estudo da fase 3 da Coronavac continua sem publicação em revista científica 1 ano após preprint. 2022. Available from: https://www1.folha.uol.com.br/equilibrioesaude/2022/05/estudo-da-fase-3-da-coronavac-continua-sem-publicacao-em-revista-cientifica-1-ano-apos-pre-print.shtml.

[24] Palacios R, Batista AP, Albuquerque CSN, Patiño EG, do Prado Santos J, Conde MTRP, et al. Efficacy and Safety of a COVID-19 Inactivated Vaccine in Healthcare Professionals in Brazil: The PROFISCOV Study. SSRN Journal. 2021.

[25] Fadlyana E, Rusmil K, Tarigan R, Rahmadi AR, Prodjosoewojo S, Sofiatin Y, et al. A phase III, observer-blind, randomized, placebo-controlled study of the efficacy, safety, and immunogenicity of SARS-CoV-2 inactivated vaccine in healthy adults aged 18–59 years: An interim analysis in Indonesia. Vaccine. 2021;39(44):6520–8.

[26] Kostis JB, Dobrzynski JM. Limitations of Randomized Clinical Trials. The American Journal of Cardiology. 2020;129:109–15.

[27] Evans SJW, Jewell NP. Vaccine Effectiveness Studies in the Field. New England Journal of Medicine. 2021;385(7):650–1.

[28] Ranzani OT, Hitchings MDT, Dorion M, D’Agostini TL, de Paula RC, de Paula OFP, et al. Effectiveness of the CoronaVac vaccine in older adults during a gamma variant associated epidemic of covid-19 in Brazil: test negative case-control study. BMJ. 2021;374. n2015.

[29] Jara A, Undurraga EA, González C, Paredes F, Fontecilla T, Jara G, et al. Effectiveness of an Inactivated SARS-CoV-2 Vaccine in Chile. New England Journal of Medicine. 2021;385(10):875–84.

[30] Dagan N, Barda N, Kepten E, Miron O, Perchik S, Katz MA, et al. BNT162b2 mRNA Covid-19 Vaccine in a Nationwide Mass Vaccination Setting. New England Journal of Medicine. 2021;384(15):1412–23.

[31] Hitchings MDT, Ranzani OT, Torres MSS, de Oliveira SB, Almiron M, Said R, et al. Effectiveness of CoronaVac among healthcare workers in the setting of high SARS-CoV-2 Gamma variant transmission in Manaus, Brazil: A test-negative case-control study. The Lancet Regional Health - Americas. 2021;1:100025.

[32] Cerqueira-Silva T, Andrews JR, Boaventura VS, Ranzani OT, de Araújo Oliveira V, Paixão ES, et al. Effectiveness of CoronaVac, ChAdOx1 nCoV-19, BNT162b2, and Ad26.COV2.S among individuals with previous SARS-CoV-2 infection in Brazil: a test-negative, case-control study. The Lancet Infectious Diseases. 2022;22(6):791-801.

[33] Hitchings MDT, Ranzani OT, Dorion M, D’Agostini TL, de Paula RC, de Paula OFP, et al. Effectiveness of ChAdOx1 vaccine in older adults during SARS-CoV-2 Gamma variant circulation in São Paulo. Nature Communications. 2021;12(1):6220.

[34] Dagan N, Barda N, Biron-Shental T, Makov-Assif M, Key C, Kohane IS, et al. Effectiveness of the BNT162b2 mRNA COVID-19 vaccine in pregnancy. Nature Medicine. 2021;27(10):1693–5.

[35] Barda N, Dagan N, Cohen C, Hernán MA, Lipsitch M, Kohane IS, et al. Effectiveness of a third dose of the BNT162b2 mRNA COVID-19 vaccine for preventing severe outcomes in Israel: an observational study. The Lancet. 2021;398(10316):2093–100.

[36] Magen O, Waxman JG, Makov-Assif M, Vered R, Dicker D, Hernán MA, et al. Fourth Dose of BNT162b2 mRNA Covid-19 Vaccine in a Nationwide Setting. New England Journal of Medicine. 2022;386(17):1603–14.

[37] Belongia EA, Simpson MD, King JP, Sundaram ME, Kelley NS, Osterholm MT, et al. Variable influenza vaccine effectiveness by subtype: a systematic review and meta-analysis of test-negative design studies. The Lancet Infectious Diseases. 2016;16(8):942–51.

[38] Verani JR, Baqui AH, Broome CV, Cherian T, Cohen C, Farrar JL, et al. Case-control vaccine effectiveness studies: Preparation, design, and enrollment of cases and controls. Vaccine. 2017;35(25):3295–302.

[39] Cerqueira-Silva T, de Araújo Oliveira V, Boaventura VS, Pescarini JM, Júnior JB, Machado TM, et al. Influence of age on the effectiveness and duration of protection of Vaxzevria and CoronaVac vaccines: A population-based study. The Lancet Regional Health - Americas. 2022;6:100154.

[40] Hernán MA, Robins JM. Using Big Data to Emulate a Target Trial When a Randomized Trial Is Not Available. American Journal of Epidemiology. 2016;183(8):758–64.

[41] Instituto Brasileiro de Geografia e Estatística. Accessed: 2022-09-19. Available from: https://cidades.ibge.gov.br/brasil/ce/fortaleza/panorama.

[42] Secretaria Municipal da Saúde de Fortaleza. Plano Municipal de Operacionalização da Vacinação Contra a Covid-19. Fortaleza, BR; 2021.

[43] IntegraSUS: Transparência da Saúde no Ceará. Accessed: 2022-09-19. Available from: https://integrasus.saude.ce.gov.br/.

[44] Staton EA. The Human Development Index: A History. 2007. Available from: https://scholarworks.umass.edu/peri_workingpapers/85/.

[45] Matthews JNS. Introduction to Randomized Controlled Clinical Trials. London, UK: Taylor & Francis; 2006.

[46] Bonanad C, García-Blas S, Tarazona-Santabalbina F, Sanchis J, Bertomeu-González V, Fácila L, et al. The Effect of Age on Mortality in Patients With Covid-19: A Meta-Analysis With 611,583 Subjects. Journal of the American Medical Directors Association. 2020;21(7):915–918.

[47] UN-HABITAT. State of the world’s cities 2010/2011: bridging the urban divide. London, UK: Earthscan; 2010.

48. Coronavírus Ceará. Passaporte de vacinação é instituído como obrigatório para eventos, bares e restaurantes. 2021. Available from: https://coronavirus.ceara.gov.br/project/passaporte-de-vacinacao-e-instituido-como-obrigatorio-para-eventos-bares-e-restaurantes/.

[49] Victora C, Castro MC, Gurzenda S, Medeiros AC, França GVA, Barros AJD. Estimating the early impact of vaccination against COVID-19 on deaths among elderly people in Brazil: Analyses of routinely-collected data on vaccine coverage and mortality. EClinicalMedicine. 2021;38:101036.

