## Supplementary Material for "Impact of CoronaVac on Covid-19 outcomes of elderly adults in a large and socially unequal Brazilian city: A target trial emulation study"

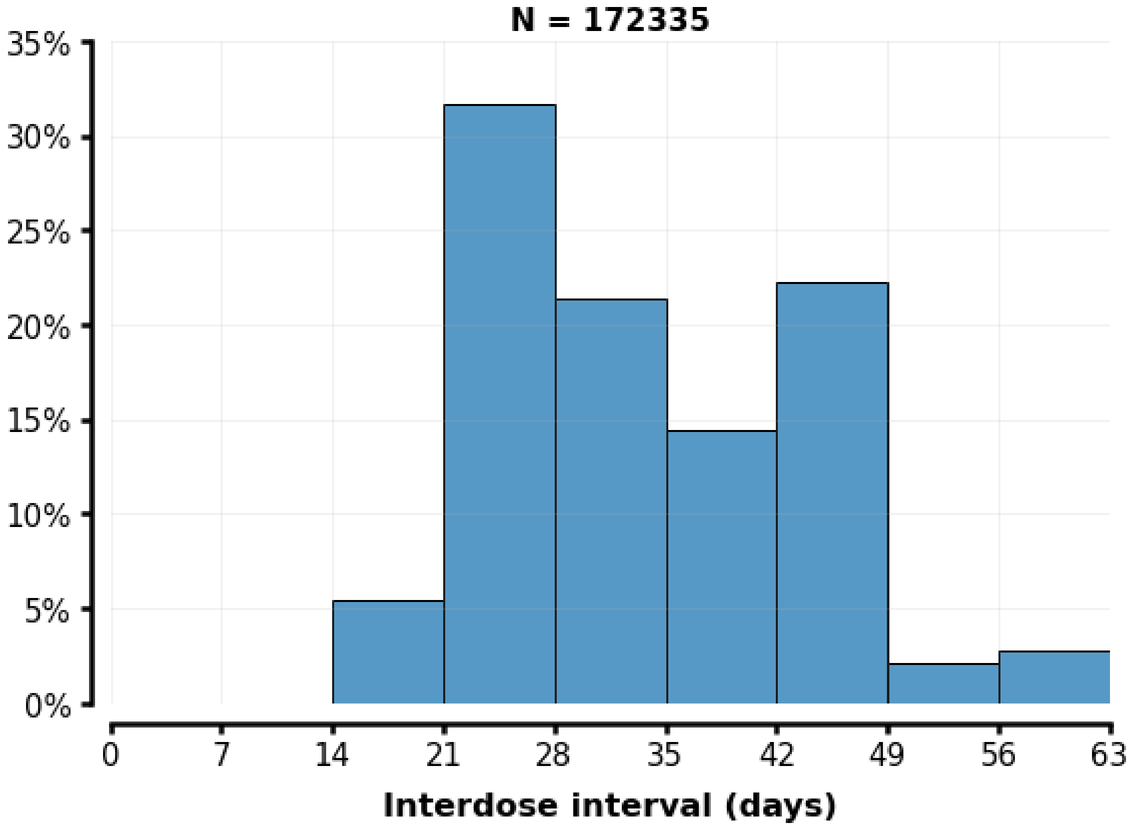


**Figure S1:** Interval between first and second dose for individuals aged ≥ 60 who received CoronaVac. During the vaccination campaign in Fortaleza, imperfect adherence to the vaccine schedule was observed, with a high percentage of individuals receiving the second dose of CoronaVac after the recommended date (maximum of four weeks).

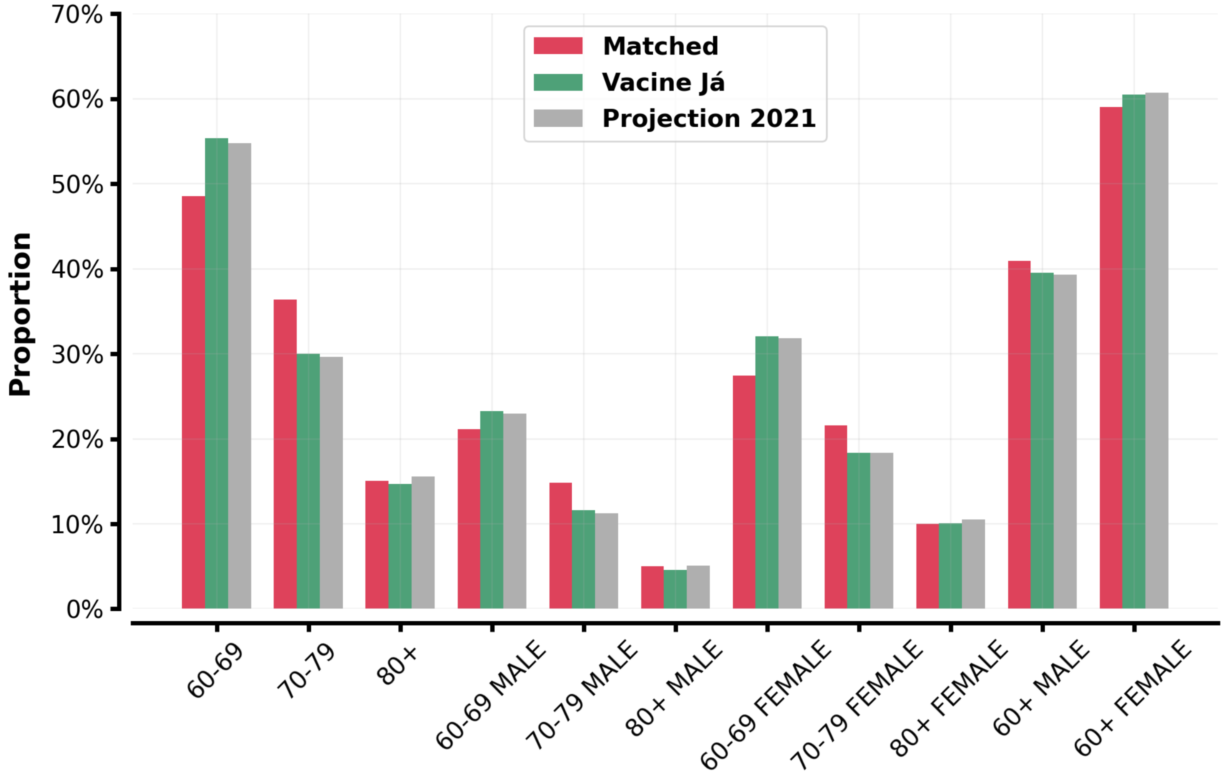


**Figure S2:** Demographic characteristics of cohort individuals, VACINE JÁ registry, and projected population for Fortaleza in 2021.

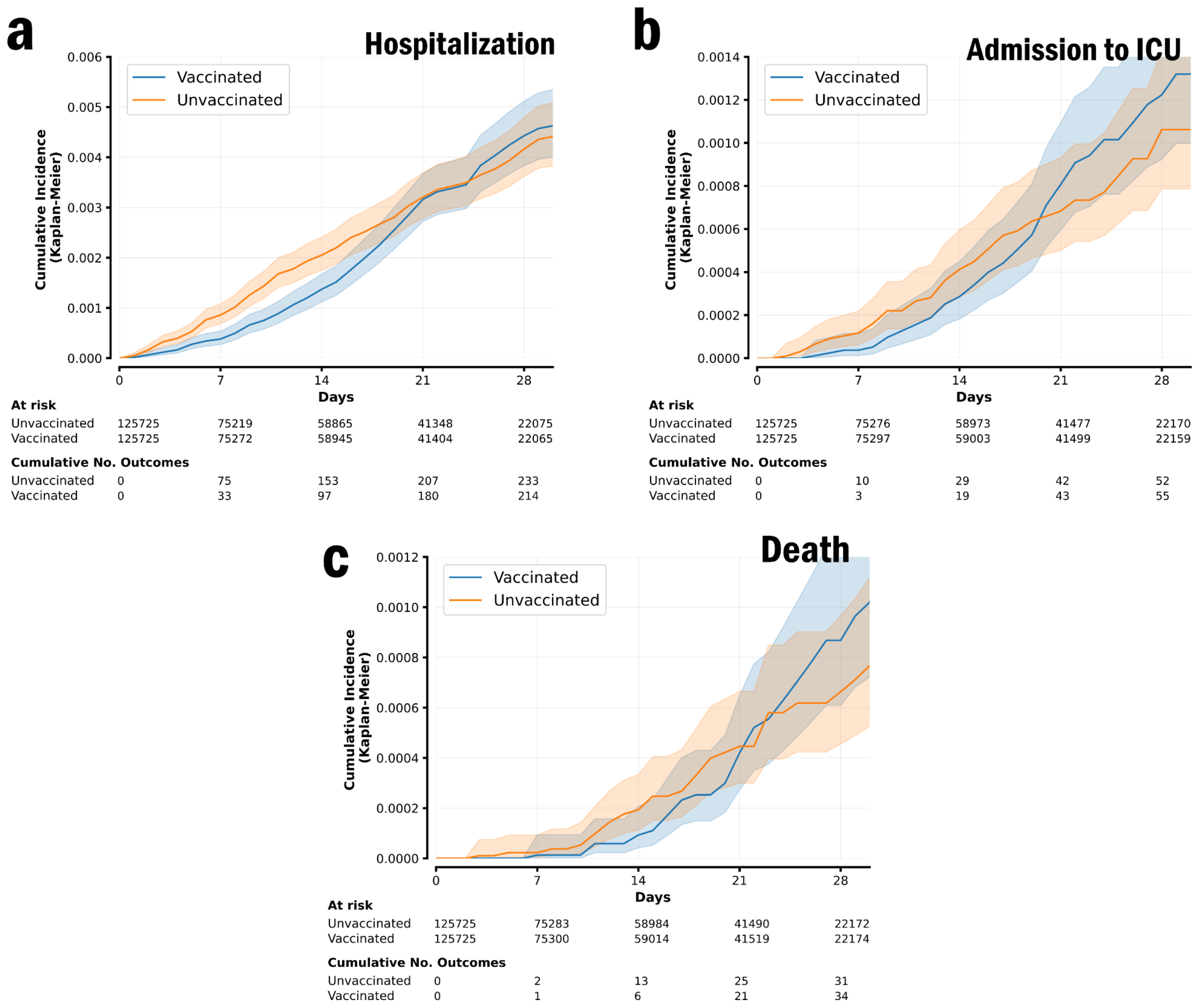


**Figure S3:** Cumulative incidence curves comparing the risk between individuals vaccinated with a single dose of CoronaVac and unvaccinated individuals. Curves were estimated with the Kaplan-Meier estimator for (a) hospital admission, (b) ICU admission, and (c) Covid-19-related death.

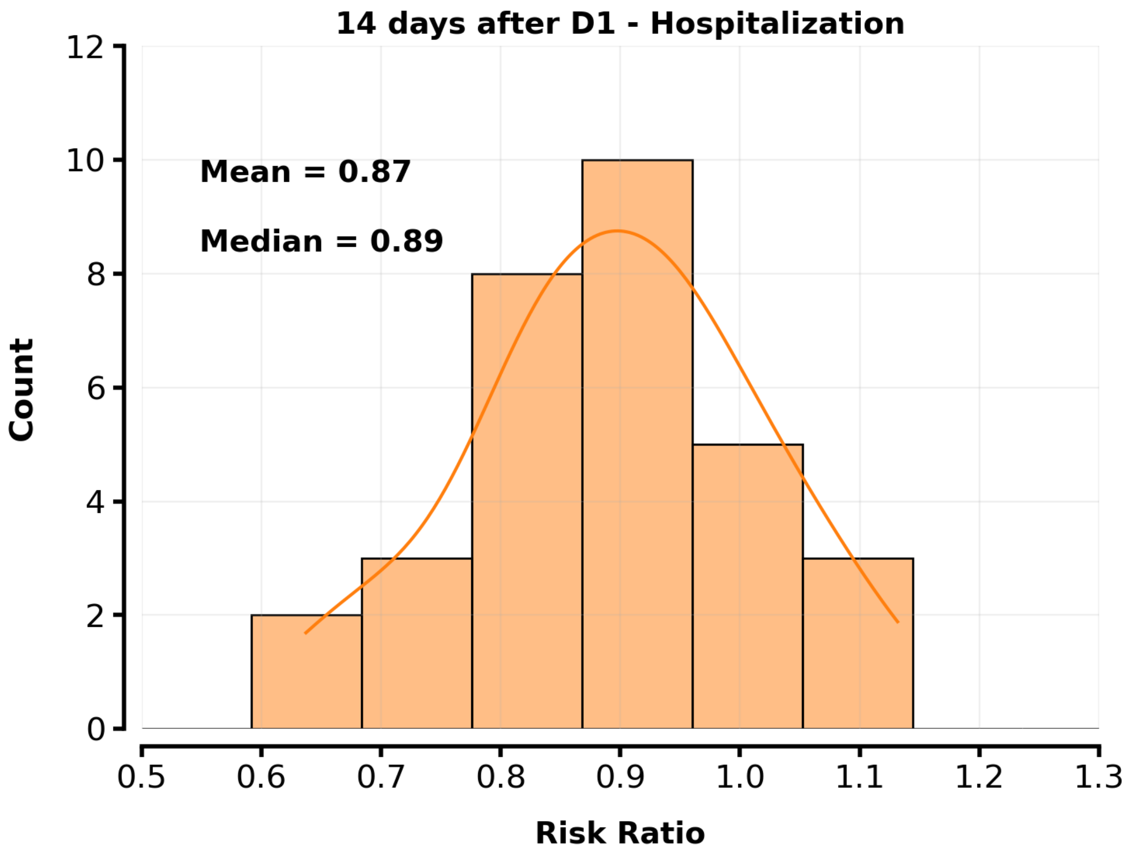


**Figure S4:** Distribution of the risk ratio between the Kaplan-Meier curves for hospitalization at day 14 after the receipt of the first dose of the vaccinated person. The distribution was obtained by using the estimated risk ratio for 30 different configurations of unvaccinated controls. The average risk ratio, estimated as 0.87 and with a standard deviation of 0.15, indicates that matched pairs have no detectable difference in their risk in the first days after the application of the first dose in the vaccinated person. Such a result corroborates the assumption of exchangeability of the matching procedure.

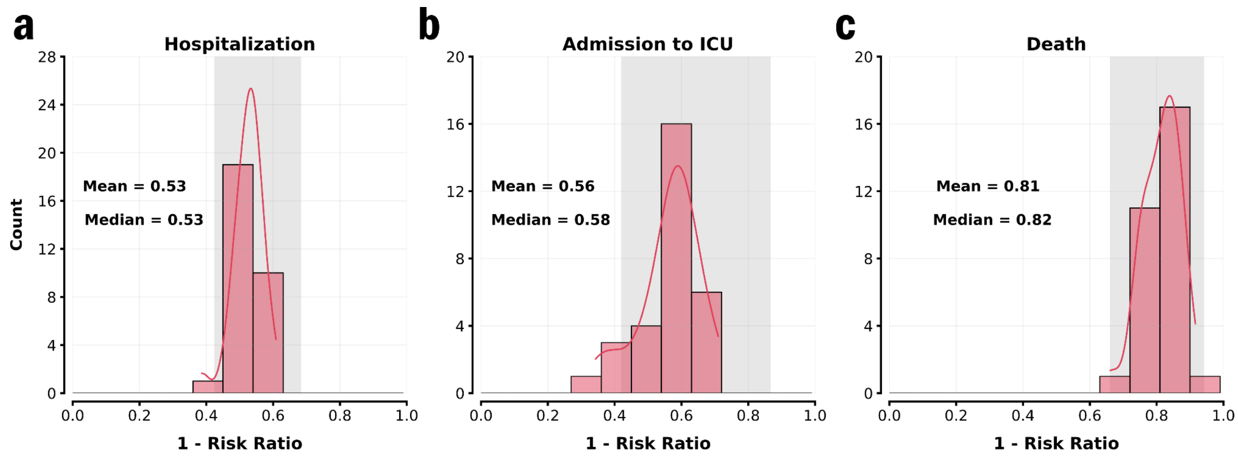


**Figure S5:** Distribution of the vaccine effectiveness (one minus the risk ratio) for the two-dose regimen of CoronaVac. (a) Hospital admission due to Covid-19 infection, (b) Severe disease manifestation through admission to ICU, and (c) Covid-19-related death. Gray area corresponds to the 95% confidence interval for each outcome in the cohort used in the main text. A total of 30 different configurations of unvaccinated controls were used to obtain the distributions.

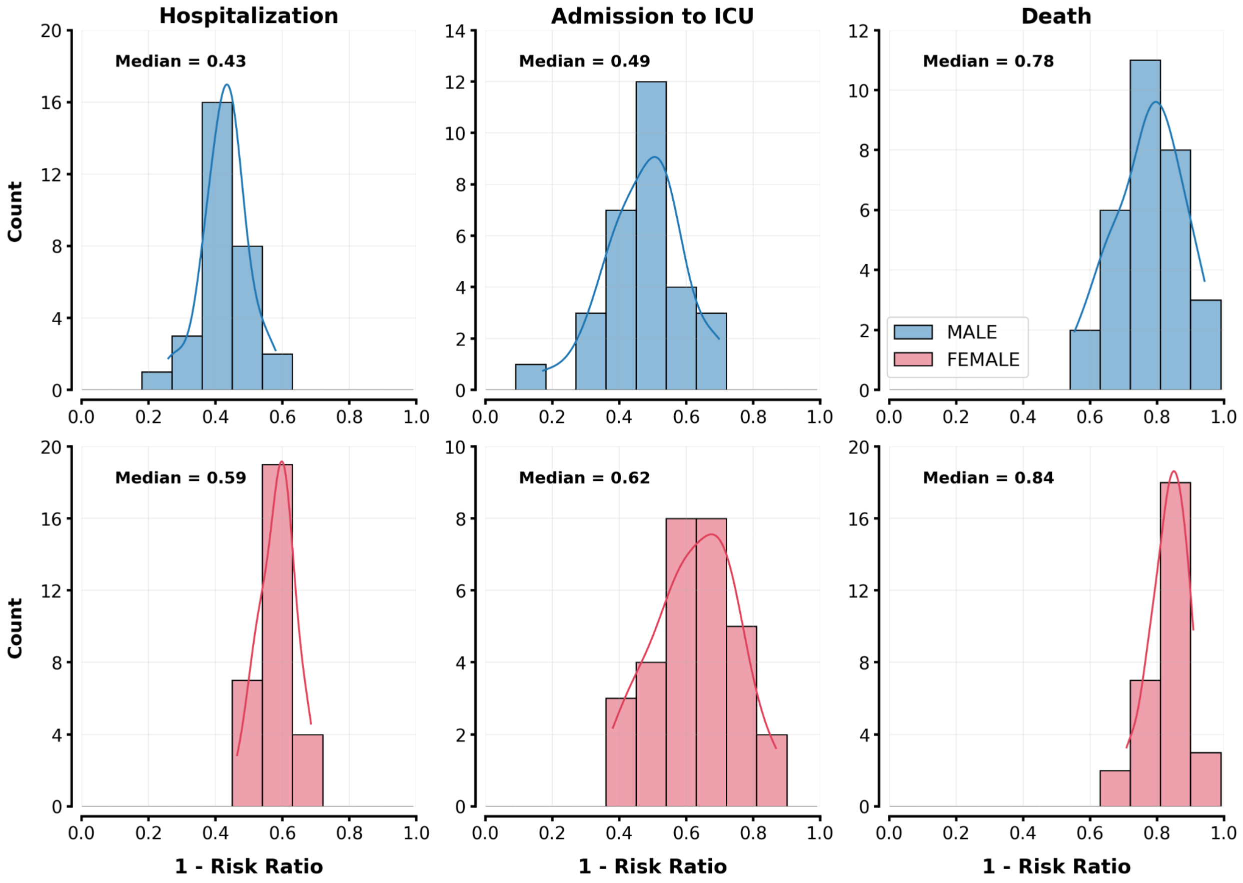


**Table S6:** Distribution of the vaccine effectiveness (one minus the risk ratio) for the two-dose regimen of CoronaVac stratified by sex. A total of 30 different configurations of unvaccinated controls were used to obtain the distributions.

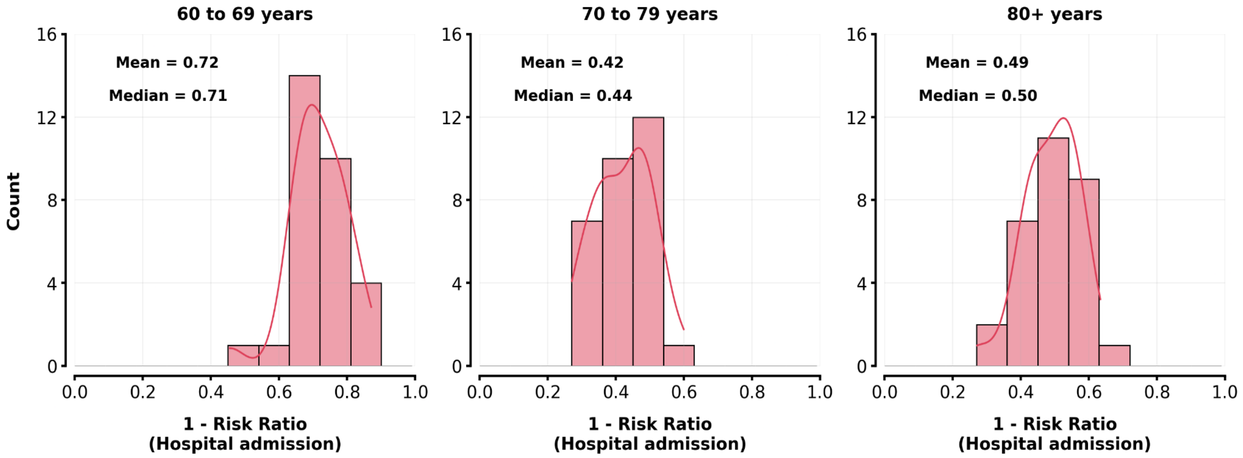


**Table S7:** Distribution of the vaccine effectiveness (one minus the risk ratio) for the two-dose regimen of CoronaVac stratified by age groups. Only the outcome of hospital admission was considered for this analysis as stratification by age produces different sample sizes of each subgroup affecting the quality of estimates for the other outcomes. A total of 30 different configurations of unvaccinated controls were used to obtain the distributions.

**Table S1:** Effectiveness of the two-dose regimen of CoronaVac against severe outcomes considering delayed censoring for pairs where the control receives a vaccine dose. We consider two scenarios: censoring is performed after 6 and 13 days after the dose receipt. For all outcomes considered, estimates of effectiveness do not exhibit significant differences between different delays and no delay. 95% confidence intervals of the estimates were obtained by using the percentile bootstrap method with 1000 repetitions.

| **Censoring** | **Outcome** | **Unvaccinated Individuals** | | **Two-dose regimen Individuals** | | **1 - RR** |
|---|---|---|---|---|---|---|
| | | **Events** | **Risk per 100,000 individuals** | **Events** | **Risk per 100,000 individuals** | **(95% CI)** |
| After 6 days | Hospitalization | 139 | 572.5 | 62 | 253.9 | 55.8% (42.3 - 68.0) |
| | Admission to ICU | 33 | 135.4 | 11 | 45.0 | 66.7% (39.5 - 84.6) |
| | Death | 47 | 193.0 | 8 | 32.7 | 82.6% (67.5 - 94.0) |
| After 13 days | Hospitalization | 153 | 606.2 | 69 | 271.9 | 54.6% (42.0 - 66.6) |
| | Admission to ICU | 36 | 142.1 | 12 | 47.2 | 66.8% (39.9 - 84.4) |
| | Death | 52 | 205.5 | 10 | 39.3 | 80.4% (64.8 - 92.5) |
